# Prevalence of SARS-CoV-2 among high-risk populations in Lomé (Togo) in 2020

**DOI:** 10.1101/2020.08.07.20163840

**Authors:** Wemboo Afiwa Halatoko, Yao Rodion Konu, Fifonsi Adjidossi Gbeasor-Komlanvi, Arnold Junior Sadio, Martin Kouame Tchankoni, Koffi Segbeaya Komlanvi, Mounerou Salou, Ameyo Monique Dorkenoo, Issaka Maman, Amétépé Agbobli, Majesté Ihou Wateba, Komi Séraphin Adjoh, Edem Goeh Akue, Yem-bla Kao, Innocent Kpeto, Paul Pana, Rebecca Kinde-Sossou, Agbeko Tamekloe, Josée Nayo-Apetsianyi, Simon-Pierre Hamadi Assane, Mireille Prince-David, Sossinou Marcel Awoussi, Mohaman Djibril, Moustafa Mijiyawa, Anoumou Claver Dagnra, Didier Koumavi Ekouevi

**Author notes:** Corresponding author Mail (YRK). Contributed equally to this work and share primary authorship (Wemboo Afiwa Halatoko, Yao Rodion Konu). Contributed equally to this work and share senior authorship (Anoumou Claver Dagnra, Didier Koumavi Ekouevi). **Authors details** WAH, YRK, FAGK, AJS, MKT, KSK, MS, AMD, IM, AA, MIW, KSA, EGA, YK, IK, PP, RKS, AT, JNA, SHA, MPD, SMA, MD, MM, ACD, DKE.

## Abstract

**Objective:** This survey aims at estimating the prevalence of SARS-CoV-2 in high risk populations in Lomé.

**Methods:** From April 23^rd^ to May 8^th^ 2020, we recruited a random sample of participants from five sectors: healthcare, air transport, police, road transport and informal. We collected oropharyngeal swab for direct detection through real time reverse transcription polymerase chain reaction (rRT-PCR), and blood for antibodies detection by serological tests. The overall prevalence (current and past) of infection was defined by positivity for both tests.

**Results:** A total of 955 participants with a median age of 36 (IQR 32-43) were included and 71.6% (n=684) were men. Around 22.1% (n=212) were from the air transport sector, 20.5% (n=196) in the police, and 38.7% (n=370) in the health sector. Seven participants (0.7%, 95% CI: 0.3-1.6%) had a positive rRT-PCR at the time of recruitment and nine (0.9%, 95% CI: 0.4-1.8%) were seropositive for IgM or IgG against SARS-CoV-2. We found an overall prevalence of 1.6% (n=15), 95% CI: 0.9-2.6%.

**Conclusion:** The prevalence of the SARS-CoV-2 infection among high-risk populations in Lomé was relatively low and could be explained by the various measures taken by the Togolese government. Therefore, we recommend targeted screening.

## Introduction

In December 2019, an outbreak of pneumonia (Covid-19) due to a new coronavirus first named 2019-nCoV, now officially SARS-CoV-2, occurred in China [1]. In less than five months, this outbreak had spread rapidly to every continent (except Antarctica) with more than 3.7 million people infected and more than 257,000 deaths recorded as of May 8^th^ 2020 in 214 countries and territories [2]. In Africa, 32,953 (0.9%) cases of Covid-19 had been reported as of May 8^th^ 2020 [3].

Since the beginning of the outbreak, health systems in developed countries have faced many challenges to fight the Covid-19. Numerous assumptions have been made about the true magnitude and evolution of the epidemic around the world. It has been commonly assumed that officially reported data are underestimated [4,5], especially in Africa. Insufficient diagnostic capacity of countries and the high proportion of asymptomatic cases may explain such underestimation [6]. Thus, the World Health Organization (WHO) has recommended a mass screening strategy for all countries burdened by the epidemic with the hypothesis that [7] more tests performed would result in an easier tracking of the spread of the virus and thus a decrease in transmission [8]. However, there is insufficient testing capacity in many countries due to a high global demand for antibody test kits [8] and GeneXpert which has recently been validated by the US Food and Drug Administration [9]. To date, real-time reverse transcription-polymerase chain reaction (rRT-PCR) remains the gold standard test for the diagnosis of Covid-19. Antibodies are the best biomarkers to estimate the number of people previously infected: its use could help estimate the prevalence and inform testing strategies in populations at higher risk of Covid-19.

In Togo, the first case of Covid-19 was reported on March 5^th^ 2020 and as of April 26^th^ 2020, 98 cases were confirmed, including 6 deaths [10]. Only suspect cases, contacts, and travellers were being screened for SARS-CoV-2. The value of population mass screening was debated considering the country’s relatively limited diagnostic capabilities. Few studies so far have been conducted to estimate the prevalence of SARS-CoV-2 based on rRT-PCR test or antibody test including studies in Iceland [11], Santa-Clara County in USA [12] and Switzerland [13]. To our knowledge, there is no data available on the prevalence of SARS-CoV-2 in Sub-Saharan Africa. Based on the low incidence of SARS-CoV-2 infection observed in the general population, the Swiss National Covid-19 Science Task Force recommends focusing research at the population level on subpopulations at higher risk of infection [14]. Therefore, we conducted a pilot survey in high risk populations to estimate the prevalence of SARS-CoV-2 using a rRT-PCR test in order to refine screening strategies in the fight against the pandemic in Togo.

## Methods

### Study design and sampling

A cross-sectional study was conducted by a multi-disciplinary team (demographers, epidemiologists, biologists, biostatisticians) among high-risk populations in Lomé (capital city of Togo) from April 23^rd^ to May 8^th^, 2020. Participants were recruited from five professional sectors: healthcare (doctors, nurses, pharmacy auxiliaries, hospital administrators), air transport, police, road transport (taxi and moto-taxi drivers) and informal (market sellers and craftsmen). These groups were targeted because they are at high risk of contamination during epidemics for their high probability of being in close contact with travelers or with Covid-19 patients [15,16]. Participants were eligible to participate in the study if the following four criteria were fulfilled: (i) ≥ 18 years of age; (ii) working in one of the five sectors; (iii) having been regularly present at the workstation for the past 30 days; (iv) living in Lomé for the past 3 months.

Several sampling methods were used for participants’ selection based on the expected total size of the target population and the availability of a sampling frame. First, exhaustive recruitment was performed among the police (road safety officers) and people in air transport *(International Airport Gnassingbe Eyadema, Lomé, Togo)*. Second, participants from the informal sector were recruited based on an open invitation. Third, random sampling (two or three stage) was performed for the recruitment of taxi motos (road transport) and health care workers. For example, for the selection of taxi moto drivers we performed a two-stage sampling with the selection of the company, then the selection of the drivers working in the company.

### Sample size estimation

The sample size was estimated using a single proportion population formula with a 95% confidence level, 1% margin of error, and 2% estimated prevalence of SARS-CoV-2 among high risk populations (as defined above) based on surveillance data in travelers in Togo (Ministry of health). A 10% unusable biological specimens or non-response rate was anticipated and the minimum number of participants was estimated at 837.

### Data collection

We established a test site at the ‘Faculté des Sciences de la Santé de l’Université de Lomé’ (Faculty of medicine, University of Lomé) and invited the target population to join us on site for inclusion. After eligibility screening and written informed consent approval, sociodemographic characteristics and Covid-19 epidemiological data were collected using a standardized questionnaire. The questionnaire was administered by a trained study team (medical doctors) during a face-to-face interview. Oropharyngeal (OP) and blood samples were collected by trained and well-equipped staff of the ‘Institut National d’Hygiène’ (INH) which is the reference laboratory for SARS-CoV-2 testing in Togo. Oropharyngeal swabs were collected using Eswab type swabs and samples were tested for SARS-CoV-2 using rRT-PCR at the INH. Whole blood specimens were collected in EDTA tubes to test for anti-SARS-CoV-2 serologic markers at the ‘Laboratoire de Biologie Moléculaire et d’Immunologie, Université de Lomé’ (BIOLIM).

### Laboratory procedures

#### Molecular detection of SARS-CoV-2 using TIB MOL BIOL rt RT-PCR Kit

Detection of SARS-CoV-2 in oropharyngeal samples has been performed using molecular biology methods, such as recommended by WHO and US CDC [17,18]. The TIB MOLBIOL (Olfert Landt, Berlin Germany) LightMix® SarbecoV E-gene plus EAV control PCR kit and the LightMix® Modular COVID-19 RdRPgene has been used for the amplification and qualitative detection of nucleic acid of SARS-CoV-2. Amplification was carried out after viral RNA extraction using the QIAamp Viral RNA Mini Kit (Qiagen Str, Hilden, Germany). The detection algorithm used was a two-step process involving first a screening assay for sarbecovirus by targeting the E gene to detect both SARS virus and COVID-19 virus; and secondly a confirmation assay for COVID-19 virus only by targeting the RNA dependent RNA polymerase gene (RdRp), specific gene for SARS-CoV-2.

Internal quality control was ensured by the use of three controls included in the kit supplied by the company TIB MOL BIOL (Eresburgstr. 22-23 | D-12103 Berlin, Germany). A control during extraction (EAV extraction control, ref. 40-0776-96, TIB MOL BIOL, Germany) to detect possible inhibition of PCR; a positive control for each gene to be detected (positive control of the E gene, ref. 40-0776-96, TIB MOL BIOL, Germany and positive control of the RdRp gene; ref. 53-0777-96, TIB MOL BIOL, Germany) to ensure good performance of the PCR reaction and a negative control (no control model, NTC) to verify the absence of contamination of the reagents. The performance of the test is only accepted if the values of the threshold cycles (Ct) of the positive controls (PTC) of E positive gene control <30; Positive control of RdRp gene <30; Control of the extraction of the EAV <33 and the NTC does not generate any amplification curve.

A sample was considered positive if the Ct genes of E <36 and RdRp <40 with the presence or absence of EAV extraction control. If the RdRp gene did not appear or the Ct> 40, the sample was considered probable at Covid-19. A sample was considered negative (below the threshold) if no amplification curve was observed for the E and RdRp genes and the EAV extraction control <33. If, on the other hand, the EAV had no curve, the test was retaliated for the corresponding sample.

The INH molecular biology laboratory participated in May 2020 in the external quality control under the supervision of WHO within the framework of its pilot program external quality assessment program (EQAP) for the detection of SARS-CoV-2 virus by RT-PCR.

#### Detection of SARS-CoV-2 antibodies

Detection of serological markers (Immunogobulins G and M) of SARS-CoV-2 infection was carried out at BIOLIM using the Lungene® Rapid Test (Hangzhou Clongene Biotech Co, Ltd). The sensitivity and specificity were 72.85% and 85.02%, respectively [19].

This test was also validated by Laboratory Department of the ministry of health in Togo. Sensitivity of the assay using samples from participants previously diagnosed with Covid-19 at the day of hospitalization was 77.1% for IgM or IgG and specificity of 95.4%. Moreover, the sensitivity was measured 7 days after hospitalization and reported a sensitivity of 93.3% for IgG signing the contact with SARS-CoV-2 [20].

### Care and treatment

Biological samples tests results were available within 48 hours. All participants screened positive for SARS-CoV-2 were quarantined in a dedicated hotel or at the national Covid-19 treatment center and those who were tested negative were invited to respect all the mitigation measures proposed by the government.

### Statistical analysis

Descriptive statistics were performed and results are presented with frequency tabulations and percentages for categorical variables. Quantitative variables are presented as medians with their interquartile range (IQR). Seroprevalence of antibodies against SARS-CoV-2, prevalence of SARS-CoV-2 infection by rRT-PCR and overall prevalence of past or current infection (positive rRT-PCR or antibody seropositivity) were estimated with their 95% confidence interval (95%CI). Comparisons of categorical variables were performed using chi-square or Fisher’s exact tests. Data analyses were performed using R© version 3.4.3 software and the level of significance was set at 5%.

### Ethical considerations

Ethical approval was obtained from the ‘Comité de Bioéthique de Recherche en Santé’ (Bioethics Committee for Health Research) from the Togo Ministry of Health (No. 004/2020/CBRS). Potential participants were informed about the study purpose and procedures, potential risks and protections. Those willing to participate were invited to sign a consent prior to participation.

## Results

### Sociodemographic characteristics

A total of 955 people with a median age of 36 (IQR 32–43) were included in the study and 71.6% (n=684) were men. Approximately 22.1% (n=212) were in the field of air transport, 20.5% (n=196) in the police, 5.8% (n=55) in the informal sector, 38.7% (n=370) in the health sector and 12.8% (n=122) in the road transport sector. None of the participants had been previously diagnosed Covid-19 positive or hospitalized in the last 30 days before the enrolment. The majority of participants, (n=936, 98.0%) were Togolese, around two-thirds (n=636, 66.6%) were in a couple and half (n=487, 51.0%) of them had university level degree. The sociodemographic characteristics according to the sector of activity are summarized in Table 1.

**Table 1:** Sociodemographic characteristics according to sector of activity, Lomé, Togo.

|  | **Air**<br>**transport** | **Police** | **Informal**<br>**sector** | **Health**<br>**sector** | **Road**<br>**transport** | **Total** |
| --- | --- | --- | --- | --- | --- | --- |
|  | **n=212** | **n=196** | **n=55** | **n=370** | **n=122** | **N=955** |
| **Age (years)** |  |  |  |  |  |  |
| <36 | 90 (42.5) | 121 (61.7) | 17 (30.9) | 158 (42.7) | 48 (39.3) | 434 (45.4) |
| ≥36 | 122 (57.5) | 75 (38.3) | 38 (69.1) | 212 (57.3) | 74 (60.7) | 521 (54.6) |
| **Sex** |  |  |  |  |  |  |
| Men | 179 (84.4) | 168 (85.7) | 34 (61.8) | 181 (48.9) | 122 (100.0) | 684 (71.6) |
| Women | 33 (15.6) | 28 (14.3) | 21 (38.2) | 189 (51.1) | 0 (0.0) | 271 (28.4) |
| **Nationality** |  |  |  |  |  |  |
| Togolese | 205 (96.7) | 196 (100.0) | 52 (94.5) | 364 (98.4) | 119 (97.5) | 936 (98.0) |
| Others | 7 (3.3) | 0 (0.0) | 3 (5.5) | 6 (1.6) | 3 (2.5) | 19 (2.0) |
| **In couple** |  |  |  |  |  |  |
| No | 49 (23.1) | 49 (25.0) | 10 (18.2) | 169 (45.7) | 42 (34.4) | 319 (33.4) |
| Yes | 163 (76.9) | 147 (75.0) | 45 (81.8) | 201 (54.3) | 80 (65.6) | 636 (66.6) |
| **Education**<br>**level** |  |  |  |  |  |  |
| None | 3 (1.4) | 0 (0.0) | 5 (9.1) | 5 (1.4) | 11 (9.0) | 24 (2.5) |
| Primary | 2 (0.9) | 1 (0.5) | 4 (7.3) | 11 (3.0) | 26 (21.3) | 44 (4.6) |
| Secondary | 105 (49.5) | 143 (73.0) | 15 (27.3) | 69 (18.6) | 68 (55.7) | 400 (41.9) |
| University | 102 (48.1) | 52 (26.5) | 31 (56.4) | 285 (77.0) | 17 (13.9) | 487 (51.0) |

Participants came from all the five health districts of the city of Lomé as shown in Figure 1.

**Figure 1:**
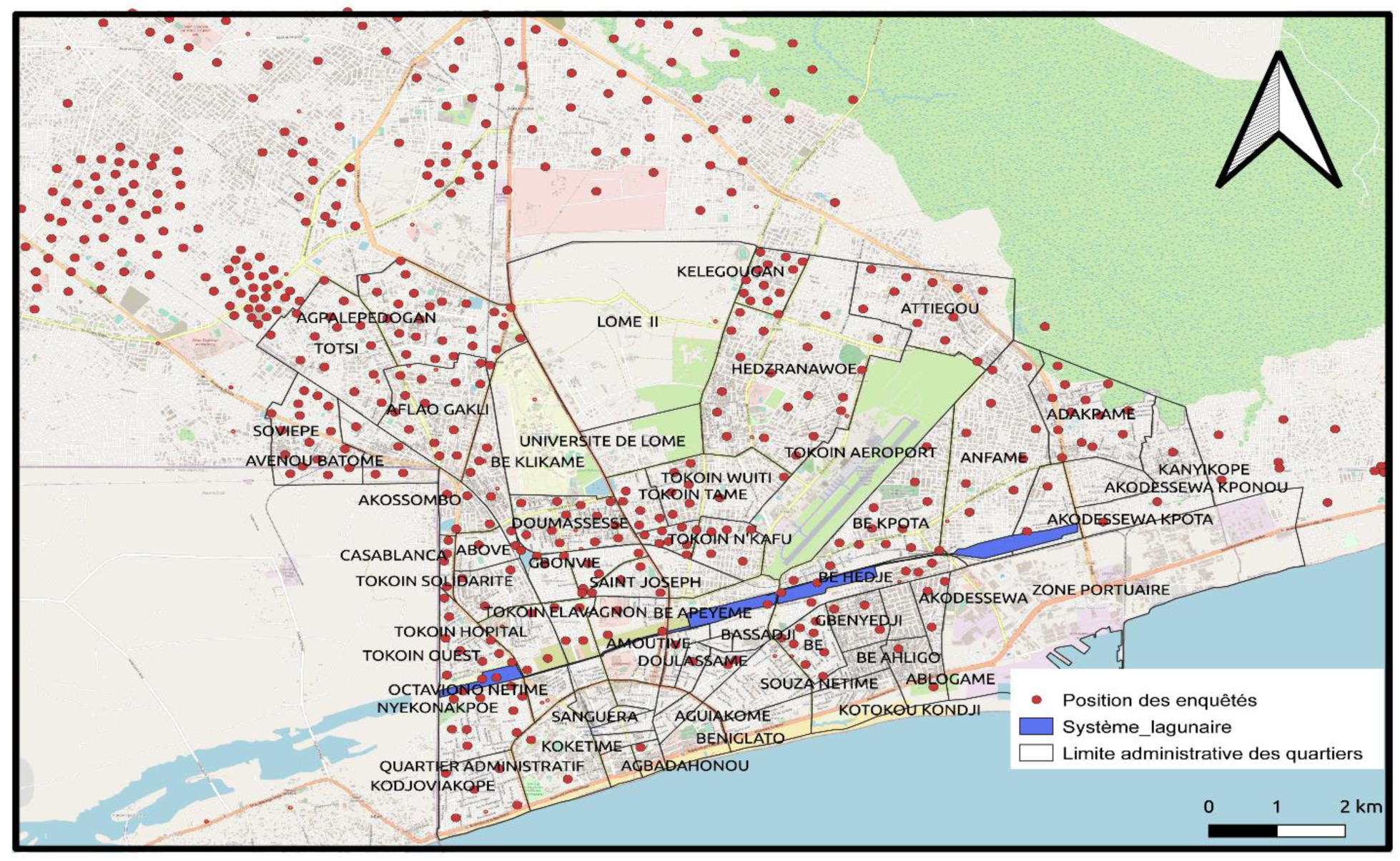
Localization of the participants in the city of Lomé (Togo)

### Prevalence of current infection as determined by a positive rRT-PCR

Seven participants (0.7%, 95% CI: 0.3-1.6%) had a positive rRT-PCR of SARS-CoV-2 at the time of recruitment and the prevalence varied from 0% for the participants from the road transport to 1.8% for those in the informal sector of activities. The prevalence was 0.7%, 95% CI [0.3-1.8] in men and 0,7%, 95% CI [0.1-2.8] in women (p=0.683).

### Seroprevalence of antibodies against SARS-CoV-2

Nine participants (0.9%, 95% CI: 0.4-1.8%) were seropositive for IgM or IgG against SARS-CoV-2 (**Table 2 and Figure 2**) and one of them was seropositive for both IgM and IgG. Table 2 summarize the seroprevalence of IgG or IgM according to the sector of activities. Thus, a total of 15 participants (1.6%, 95% CI: 0.9-2.6%) were positive for rRT-PCR or seropositive for IgM or IgG against SARS-CoV-2. This prevalence ranged between 0.8% in the road transport sector and 1.9% in the health sector (Table 2).

**Figure 2:**
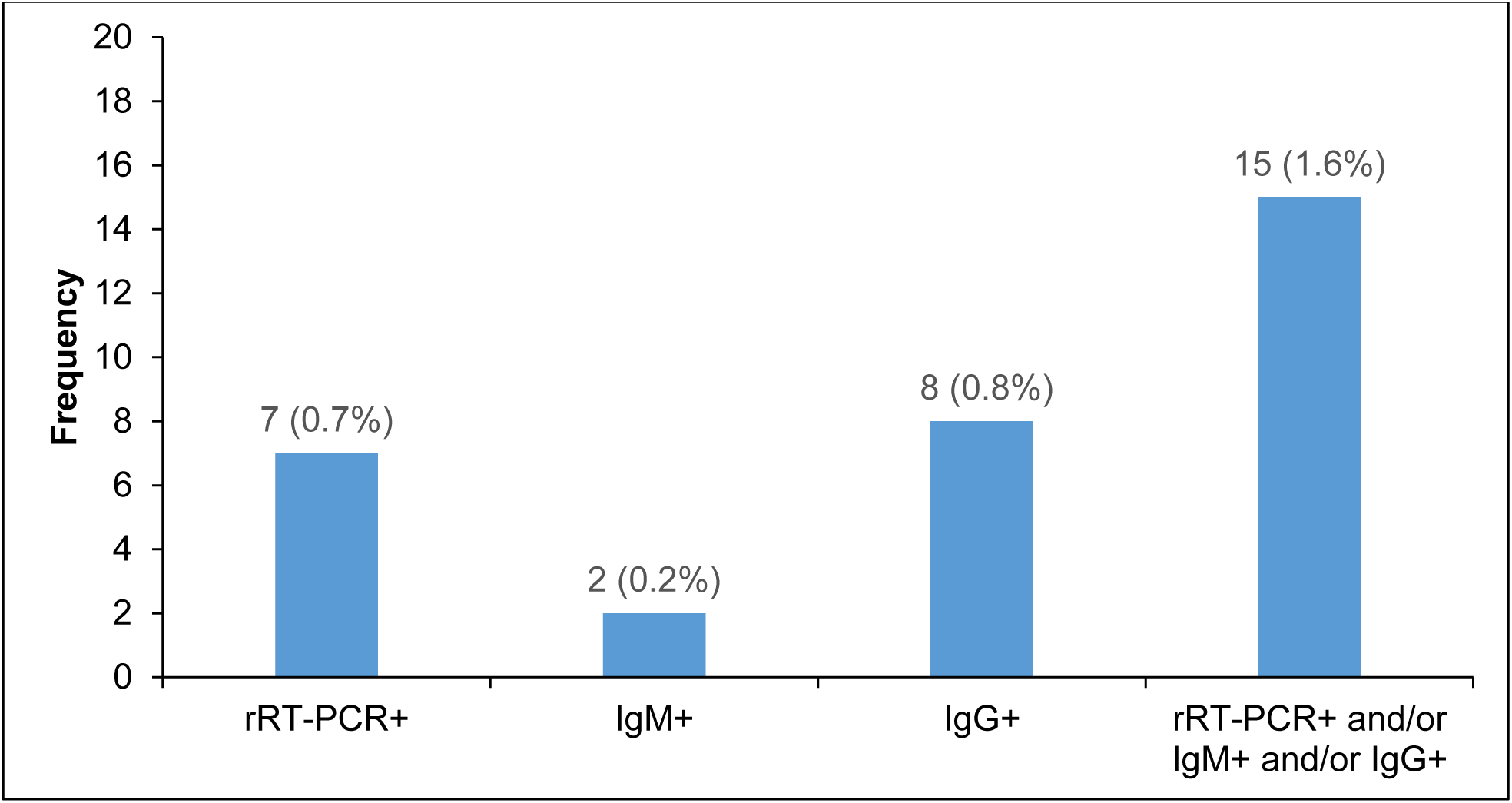
Prevalence of SARS-CoV-2 based on rRT-PCR test and serological test in high risk population, Lomé (Togo).

**Table 2:** Prevalence of SARS-Cov-2 according to sector of activity in Lomé, Togo.

|  | Air<br>transport | Police | Informal<br>sector | Health<br>sector | Road<br>transport | Total | 95%CI |
| --- | --- | --- | --- | --- | --- | --- | --- |
|  | n=212 | n=196 | n=55 | n=370 | n=122 | N=955 |  |
| **rRT-PCR+** | 3 (1.4) | 1 (0.5) | 1 (1.8) | 2 (0.5) | 0 (0.0) | 7 (0.7) | [0.3-1.6] |
| **IgM+** | 1 (0.5) | 1 (0.5) | 0 (0.0) | 0 (0.0) | 0 (0.0) | 2 (0.2) | [0.03-0.8] |
| **IgG+** | 2 (0.9) | 0 (0.0) | 0 (0.0) | 5 (1.4) | 1 (0.8) | 8 (0.8) | [0.4-1.7] |
| **rRT-PCR+ or IgG+ or IgM+** | 4 (1.9) | 2 (1.0) | 1 (1.8) | 7 (1.9) | 1 (0.8) | 15<br>(1.6) | [0.9-2.6] |

**+: Positive** **95%CI: 95% Confidence interval**

### Clinical manifestations, care and treatment

Six out of seven rRT-PCR positive participants were asymptomatic. The symptomatic participant had presented fever, headaches, and myalgia during the two weeks prior to enrolment. All rRT-PCR positive participants were hospitalized at the national reference center for Covid-19 and treated with hydroxychloroquine and azithromycin as recommended by the national guidelines [21]. Description of the seven rRT-PCR positive participants is presented in Table 3.

**Table 3:**
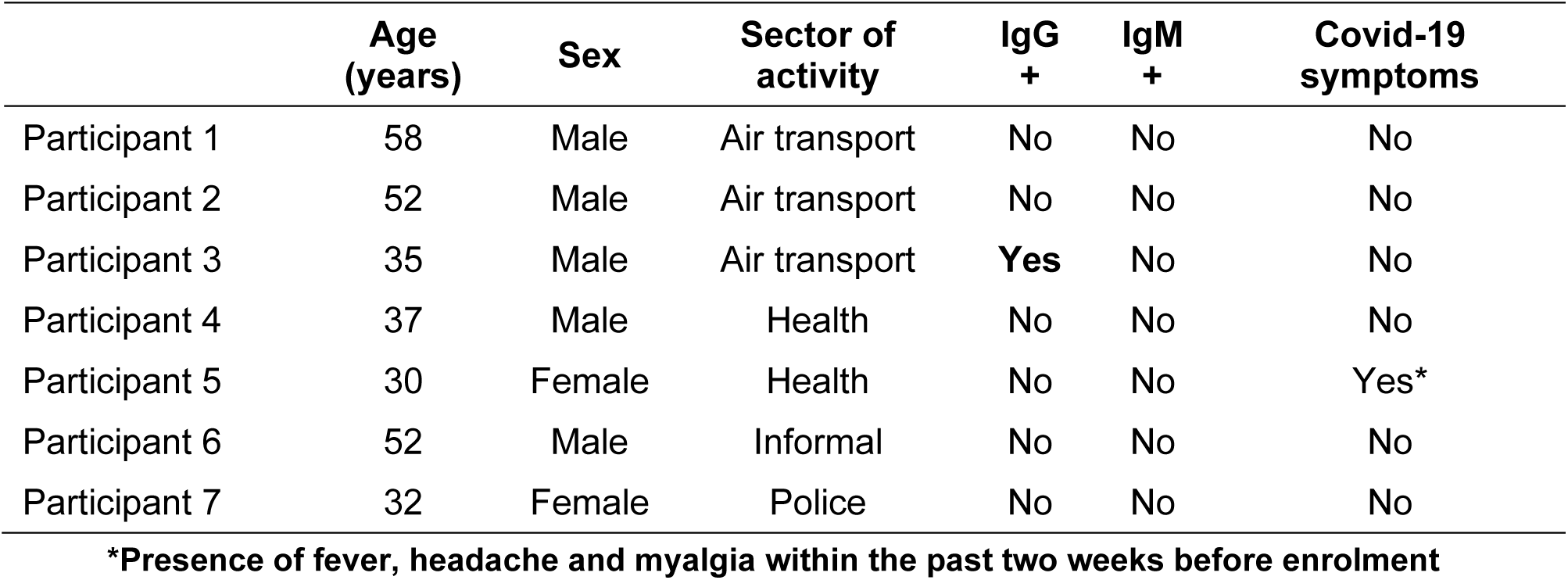
Description of rRT-PCR positive participants (N = 7)

## Discussion

To our knowledge, this is the first study reporting the prevalence of SARS-CoV-2 in sub-Saharan-Africa in a representative sample of high-risk populations. The present study was conducted in a context of urgent need for data for decision-making and refinement of response strategies. SARS-CoV-2 prevalence was assessed by molecular biology and serologic tests. In Lomé, capital city of Togo, 2 months after the first case of Covid-19, the prevalence of SARS-CoV-2 infection among persons at high risk for infection was 1.6% based on the presence of antibodies and viral genomes. Using rRT-PCR alone, only 0.7% of the study population was found to be infected with SARS-CoV-2.

Few studies have focused on high risk populations for Covid-19. In the United States, a screening of an alleged high-risk population of residents and staff members from five homeless shelters was conducted in the cities of Boston, San Francisco, Seattle and Atlanta [22]. This study reported a prevalence of SARS-CoV-2 infection ranging from 4 to 66% among residents and 1 to 30% among staff [22]. Healthcare workers (HCWs) are particularly considered as high-risk populations. In a study conducted in United Kingdom (UK), over a 3-week period (April 2020), 1,032 asymptomatic HCWs were screened for SARS-CoV-2 in a large UK teaching hospital. rRT-PCR was used to detect viral RNA from a throat and nose self-swab [23]. Among these asymptomatic HCWs, 3% were tested positive for SARS-CoV-2 [23]. In Lomé, the prevalence among HCWs was 0.5% based on virological test and 1.9% (virological and antibody test), but this included administrative and pharmacy staff.

Most Covid-19 prevalence surveys have been carried out in the general population. In a survey conducted in Iceland, 1,221 (13.3%) of the 9,199 people who were recruited using the symptom-targeted method were positive for SARS-CoV-2 infection [11]. Among those tested by open invitation selection or random selection, SARS-CoV-2 prevalence was 0.8% and 0.6%, respectively [11]. In another survey in Santa Clara County, California, USA, the crude prevalence of SARS-CoV-2 antibodies was 1.5% [12]. After weighing for population demographics of Santa Clara County, the prevalence was 2.8% [12]. In Geneva, Switzerland, another study found that the seroprevalence of SARS-CoV-2 in the general population was low (about 5%), despite the high incidence of Covid-19 in Geneva compared with other cantons [13]. All of the studies carried out in the general population based on survey or mathematical model such as in France (4.4%), reported low prevalence of SARV-CoV-2 despite the magnitude of the infection [24].

Recent evidence highlighted a highest sensitivity of the use of nasopharyngeal samples compared to oropharyngeal ones in rRT-PCR [25]. In a study conducted in China including 353 patients, using both oropharyngeal and nasopharyngeal swabs, SARS-Cov-2 rRt-PCR was positive in 19.0% of nasopharyngeal specimens against 7.6% in oropharyngeal [26]. Another survey reported that the SARS-CoV-2 detection rate was significantly higher for nasopharyngeal swabs [46.7% (56/120)] than oropharyngeal swabs [10.0% (12/120)] (P < 0.001) [25]. This could certainly contribute to underestimate the prevalence reported in our population. This prevalence could be multiplied by two or three according to available data but remain less than 3%. However, at the time of the survey there was no clear recommendation on which swab to choose. Also, unavailability of nasopharyngeal swabs didn’t allow us to collect both specimens.

The use of serological tests as an effective method for the detection of SARS-CoV-2 has been reported previously in the literature. In a study conducted in Wuhan, China, of the 56 enrolled symptomatic patients, 40 (71%) showed negative nucleic acid tests and 16 (29%) were positive. Among the 40 negative patients, 34 (85%) tested positive for the presence of IgM antibodies. Among the 16 patients who tested positive with nucleic acid tests, one patient showed a negative IgM level. The IgG antibody test was positive in all 56 patients [27]. In the present study, the prevalence of SARS-CoV-2 increased to 1.6% when serology results were included in the analysis. Based on the validation of the test in Togo and elsewhere, the data reported on serological samples seem reliable. Since serology is rapid and inexpensive it could constitute an attractive screening alternative for countries with limited resources in the context of easing of restrictive measures.

This study targeted people considered at high risk for Covid-19 based on professional activity as recently recommended by the Swiss National Covid-19 Science Task Force. The WHO also recommends to conduct survey to estimate the prevalence of SARS-CoV-2 in the community, but also for critical population subgroups such as nursing homes or health care facilities [28].

This study had some limitations. First, the population included was not representative of the general Togolese population, we actively chose this selection criterion to have a high probability of identifying cases of SARS-CoV-2 infection. Second, we cannot exclude selection bias in our sample due to the recruitment methods. Based on the low prevalence, we did not study the association between prevalence of SARS-CoV-2 according to the different sectors. Finally, this study was conducted only in Lomé, capital city of Togo, where 55% of cases of Covid-19 were identified at the time of the survey.

## Conclusion

In conclusion, the prevalence of the SARS-CoV-2 infection in the capital city among high-risk populations was relatively low two months after the notification of the first case. The low circulation of the virus in high-risk populations could be explained by the various measures taken by the Togolese government. Based on this result, generalized screening of SARS-CoV-2 would be time-consuming, not cost effective and at a high risk of reagent rupture. Therefore, we recommend targeted approach for screening. Targeted screening could include health professionals, airport staff; teachers could also be considered in the event of schools’ reopening. Repeated prevalence surveys are needed to refine the strategies to fight against Covid-19 in Togo.

## Data Availability

Data are available on request by sending an email to authors.

## List of abbreviations

95% CI: 95% confidence interval
IQR: interquartile range

## Acknowledgements

We are thankful to the participants who accepted to participate in this study and the medical students of the ‘Faculté des Sciences de la Santé-Université de Lomé’ who performed data collection for the study.

We hereby also acknowledge the work of the teams of the two laboratories involved in this study, the members of sectorial unit of management of pandemic in the ‘Ministère de la Santé et de l’Hygiène Publique’ (Ministry of Health), the rapid response teams of health region of Lomé, the staff of Santé Intégrée and the World Health Organization Representative in Togo.

Finally, we thank, Alexandra Bitty-Anderson, Abidjan, Côte d’Ivoire; El Houda Roukaya, Alex DeVille and Hannah Wang who edited the English version of this paper.

